# Effectiveness of non-pharmaceutical measures (NPIs) on COVID-19 in Europe: A systematic literature review

**DOI:** 10.1101/2021.11.11.21266216

**Authors:** Constantine I. Vardavas, Katerina Nikitara, Katerina Aslanoglou, Michele Hilton-Boon, Revati Phalkey, Jo Leonardi-Bee, Gkikas Magiorkinis, Paraskevi Katsaounou, Anastasia Pharris, Ettore Severi, Jonathan E. Suk

**Affiliations:** School of Medicine, University of Crete, Greece; Department of Oral Health Policy and Epidemiology, Harvard School of Dental Medicine, Harvard University, Boston, MA, USA; WISE Centre for Economic Justice, Glasgow Caledonian University; Division of Epidemiology and Public Health, School of Medicine, University of Nottingham, UK; Climate Change and Health Group, Public Health England, United Kingdom; Department of Hygiene, Epidemiology and Medical Statistics, Medical School, National and Kapodistrian University of Athens, Greece; Department of Respiratory Medicine, National and Kapodistrian University of Athens, Athens, Greece; Pulmonary and Respiratory Failure Department, First ICU, Evaggelismos Hospital. Athens, Greece; European Centre for Disease Prevention and Control, Solna, Sweden

## Abstract

**Background:** The study objective was to conduct a systematic review to assess the effectiveness of non-pharmaceutical interventions (NPIs) to reduce the transmission of SARS-CoV-2 in Europe during the first wave of the pandemic.

**Methods:** We searched OVID Medline, EMBASE, and the Cochrane and Campbell Databases for Systematic Reviews published up to April 15^th^ 2021. Focusing on community (meso-level) and society (macro-level) level NPIs, we included all study designs, while a geographic restriction was limited to the EU, UK and European Economic Area (EEA) countries. Using the PICO framework, two reviewers independently extracted data and assessed quality using appropriate quality appraisal tools. A qualitative synthesis was performed, with NPIs grouped initially by a) Physical Distancing measures, b) Case detection and management measures, and c) hygiene measures and subsequently by country.

**Results:** Of 17,692 studies initially assessed, 45 met all inclusion criteria. Most studies (n=30) had a modelling study design, while 13 were observational, one quasi-experimental and one experimental. Evidence from across the European continent, presented by country, indicates that the implementations of physical distancing measures (i.e., lockdowns/quarantines), preferably earlier in the pandemic, reduce the number of cases and hospitalisation across settings and for which the timing and duration are essential parameters. Case detection and management measures were also identified as effective measures at certain levels of testing and incidence, while hygiene and safety measures complemented the implementation of physical distancing measures.

**Conclusions:** This literature review represents a comprehensive assessment of the effectiveness of NPIs in Europe up to April 2021. Despite heterogeneity across studies, NPIs, as assessed within the context of this systematic review at the macro and meso level, are effective in reducing SARS-CoV-2 transmission rates and COVID-19 hospitalisation rates and deaths in the European Region and may be applied as response strategies to reduce the burden of COVID-19 in forthcoming waves.

## Introduction

Since its emergence in December 2019 in Wuhan, China, and its spread across the globe, COVID-19 has developed into a pandemic, heavily impacting society and healthcare systems. As of September 26^th^, 2021, more than 37.5 million COVID-19 confirmed cases and 760,000 deaths had been reported in the European Union/European Economic Area (EU/EEA) (1). Undoubtedly, transmission is still widespread, although stable or decreasing hospitalisation and death rates were noted in most European countries during summer period, which has led to the reduction of non-pharmaceutical interventions (NPIs). However, the emergence of SARS-CoV-2 variants with increased transmissibility, notably the B.1.617.2 Delta variant (2), the strain on healthcare systems, the economic burden and seemingly plateauing vaccination coverage in the EU, may require continued large proportions of the population that remain unvaccinated to continue to implement NPIs as a public health response to the pandemic (3, 4). It is imperative that such decisions are based upon the best available evidence while also taking into account the context in which NPIs are implemented.

NPIs aim to reduce SARS-CoV-2 transmission through physical distancing and/or through minimising the risk of social interactions that may be larger than that (5). A broad range of different NPI responses has been adopted in Europe and worldwide, mainly in the control of other respiratory virus epidemics (6). NPIs implemented across Europe are numerous and include hygiene measures such as hand hygiene and protective mask-wearing, national or international travel bans and restrictions, physical distancing measures such as school and workplace closures and gathering bans, and health system measures such as testing and contact tracing strategies.

Determining the effectiveness of NPIs in specific regions and population groups is of high priority for policymakers, who must weigh a range of options that seek to enable social activities while also preventing SARS-CoV-2 transmission. The research community has dedicated significant resources to estimate the effectiveness of NPIs. As population concerns and individual behaviours have continuously been changing throughout the COVID-19 pandemic, targeted research efforts are needed to better understand changes in the effectiveness of NPIs with time and according to specific contexts. Given the abovementioned, this systematic review aims to assess the effectiveness of NPIs implemented in Europe, primarily up to early autumn of 2020, before vaccines were available.

## Methods

### Search Results and PICO

Relevant peer-reviewed studies were identified through systematic electronic searches using OVID Medline, EMBASE and the Cochrane and Campbell Databases for Systematic Reviews. The detailed search strategies for the biomedical databases are presented in **Supplementary File 1** and included a timeframe between July 27^th^, 2020 and until April 15^th^, 2021 and were limited to the EU, UK and EEA countries, within the context of a service contract commissioned by European Center for Disease Prevention and Control (ECDC). Study designs included peer-reviewed studies including but not limited to: Simulation studies using mathematical models or transmission models, cluster and parallel randomised controlled trials (RCTs), cluster and parallel non-RCTs, quasi-experimental studies (including controlled before-after studies, uncontrolled before-after studies, time series, and interrupted time series) that assess the effectiveness of community non-pharmaceutical measures. Systematic and non-systematic literature reviews were identified, and reference lists were screened for identifying further studies.

The following set of inclusion criteria, based on the PICO framework (P-Population, I-Intervention, C-Comparison, O-Outcomes) for systematic reviews (7), was used to determine the eligibility of the studies and include: Populations were restricted to studies on humans, the assessed NPIs included community (meso-level) and society (macro-level) mitigation measures for COVID-19, containing, but not limited to, closure of educational institutions, closure of public places (mandatory and voluntary), travel bans (closure of points of entry), travel advice, personal protective equipment, and physical distancing. Excluded interventions include individual (micro-level) measures, such as the individual effect of the use of personal protective equipment. The unmitigated pandemic, baseline, or the period before NPI implementation were used as the comparator. Finally, primary outcome measures include: i) COVID-19 cases, incidence and peaks, growth rate, and ii) R as an index of transmission. Secondary outcome measures include: i) mortality associated with COVID-19, ii) Intensive Care Unit (ICU) and hospital admissions.

### Study selection

Studies identified from the searches were uploaded into a bibliographic database, and duplicates were removed. Initially, a pilot training title/abstract screening process was used, where a random sample of 100 titles was independently screened for eligibility by two reviewers to enable consistency in screening and identify areas for amendments in the inclusion criteria. A high measure of inter-rater agreement was achieved (percentage agreement>90%), and hence the remaining titles were distributed between the two reviewers and screened independently. For the full-text screening, a similar process was followed. Ten randomly selected studies were independently screened for eligibility by two reviewers (percentage agreement>90%), while the two reviewers subsequently screened the remaining full texts. Any disagreements were thoroughly discussed with a third reviewer.

### Data extraction, synthesis and presentation

Data extracted were related to the study characteristics (first author’s name, year of publication), geographical context (country/area), setting (where the measures were implemented), population characteristics, sample size, study type, numerical or descriptive findings with regards to the effectiveness of non-pharmaceutical measures in comparison to no intervention or current situation. Two reviewers independently piloted the data extraction template on a random sample of five included studies to assess consistency in data extraction and identify where amendments need to be made to the template. The remaining studies were then data extracted independently by the two reviewers.

The results from the studies for each intervention grouping are synthesised initially using a narrative synthesis. Characteristics of the included studies are also presented in the manuscript in tabulated form and provide details on the study design, geographical area, characteristics of the considered populations, setting, context, description of the studied NPIs and the qualitative and quantitative findings of the studies. The effectiveness of interventions is grouped based on the type of NPI. Areas of commonality between the results of the studies were identified through conducting a content analysis using an inductive approach, where the concepts are derived from the data. The categorisation primarily performed by the different types of NPIs is as follows:

✓ **Combined physical distancing measures:** Combinations of travel restrictions, school and university closures, closure of non-essential businesses, teleworking, closure of entertainment venues, ban on events, restrictions on mass gatherings, stay-at-home orders, shielding of older people, self-isolation, quarantine of (suspected) cases, curfew, lockdown.
✓ **School closure:** A few studies evaluated the individual effect of school closures and, hence, they are presented separately.
✓ **Case detection and management:** Contact tracing, isolation of confirmed cases and quarantine of contacts/suspected cases.
✓ **Hygiene measures:** Face mask and other protective equipment, disinfection of surfaces and air safety (from a meso or macro level).

The second level of grouping was performed by country as NPIs would be more comparable in the same setting, given that across countries, different combinations of NPIs were implemented in different timeframes and with varying levels of public compliance. Studies utilising combined data from multiple countries were also grouped together. Within the same country, studies are presented by study design, starting from the simulation studies and continuing with quasi-experimental and observational studies. As mentioned in the study of origin, the terms and descriptions of measures have been transferred verbatim to this systematic review and no new definitions have been developed. By that, lockdown includes a different combination of measures for each studied country.

### Assessment of Study Quality

Given the various study types identified in this current review, multiple quality appraisal tools were applied to assess the included studies’ quality. The Joanna Briggs Institute (JBI) standardised critical appraisal tools were used for RCTs, quasi-experimental and cohort studies (8), the Effective Practice and Organisation of Care (EPOC) Risk of Bias tool for time series (9), and for modelling studies, a tool developed by Burns et al. (2020) was utilized (10). Three studies with a time trend analysis design were not appraised due to the lack of a reliable tool.

## Results

### Study Selection

A total of 17,692 studies were identified through systematic electronic searches across Medline and Embase. After removing duplicates, 17,661 passed onto the title/abstract review process. Subsequently, 634 studies were found to meet the inclusion criteria after the completion of abstract screening and were further screened for eligibility based on full-text assessment. Through the full-text screening, 588 studies were excluded due to the reported study type (reviews, conference abstracts), little relevance to the topic, limited outcomes of interest, limited data and not eligible geographical area. Hence, 45 studies were eventually considered in our systematic review. The flowchart of study selection and exclusion is presented in **Figure 1**.

**Figure 1.**
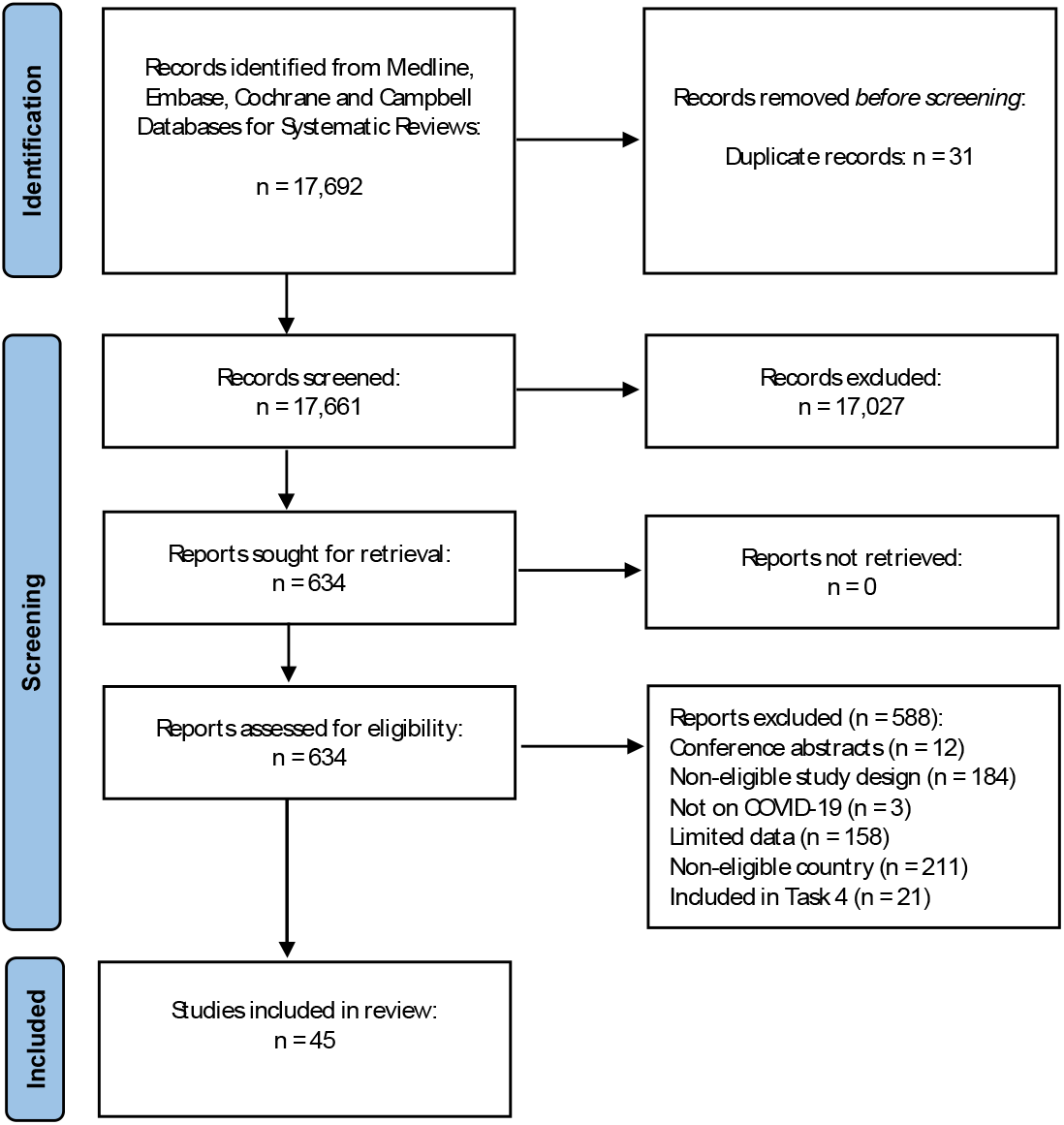
PRISMA Flowchart of the screening process.

### Study Characteristics

Among the 45 studies included in this current systematic review, 30 studies were modelling, one was an RCT, one followed a quasi-experimental approach, three were time series, three used a time-trend analysis, and seven were cohort studies. Concerning the type of NPI intervention, 40 studies evaluated different physical distancing measures, while 4 studies focused on case detection and contact tracing strategies and 3 on the implementation of hygiene measures. With regard to the geographical area, studies assessed data from Italy (n=7), Spain (n=3), England (n=9), Scotland (n=1), France (n=3), Germany (n=4), Portugal (n=2), Greece (n=1), Sweden (n=1), Denmark (n=2), Belgium (n=2), Norway (n=1) and the Netherlands (n=1), while 8 studies considered multiple European countries in their analyses.

### Quality Appraisal

Variability in study quality was noted in the 31 included modelling studies, but some patterns were also found. In several studies, there were concerns about the suitability of structural assumptions and parameters used given the multifactorial nature of interventions, their varying level of implementation and the inconsistent values published in the literature. Additionally, the investigation of uncertainty was either inadequate or not sufficiently reported in many studies. No study was excluded due to study quality; however, the results of our quality appraisal per included study are presented in detail in **Supplementary File 2**.

## Combined physical distancing measures

Overall, different combinations of physical distancing (SD) measures were investigated in terms of effectiveness in modelling and non-modelling approaches.; these are presented separately for ease of comparison due to heterogeneity in the implementation of NPIs across EU/UK/EEA countries, given the different combinations of applied measures, as well as the differences on the timeframes and the compliance of the general population, the results are presented per country to allow for internal comparisons.

### Italy

#### Modelling studies

Four modelling studies were included in this current review with data from Italy. The effectiveness of lockdown measures implemented primarily in Lombardy and 15 northern provinces on March 3^rd^, 2020, and in the whole country afterwards (March 11^th^, 2020) was examined by Gatto et al. (11). Applying an SEIR model (Susceptible-Exposed-Infectious-Recovered), the authors parametrized data from 107 provinces between February 21^st^ and March 25^th^, 2020 and suggested that the SD measures reduced transmission by 45%, while 0.226 × 10^6^ cases were averted. Palladino et al. (12) showed that a 7-day earlier introduction of a lockdown across Italy would have averted 126,000 COVID-19 cases, 12,800 deaths, 54,700 non-ICU admissions and 15,600 ICU admissions in Italy, while Marzianoa et al. (13) within a SIR (Susceptible-Infectious-Recovered) model simulated different scenarios for an earlier lifting of lockdowns, and concluding that an earlier lifting would have led to higher hospitalisation rates, as it was modelled that an earlier opening on May 4^th^ instead of May 18^th^, would lead to a twofold (+118%) increase in the incidence of hospital admissions. This effect would be even more profound if the lockdown were to be lifted even earlier, on April 20^th^, with an increase in the incidence of hospitalisations (+472%), while school closure would in parallel lead to a further reduction in cases. The investigation of the COVID-19 situation in Italy is complemented by the study of Sjodin et al. (14), who noted the significance of adherence to NPIs for the effective management of the pandemic in addition to the timing and duration.

#### Non-Modelling studies

Among the three non-modelling studies, Silverio et al. (15) found a strong positive relationship between the number of confirmed COVID-19 cases before lockdown and mortality up to 60 days later and between the daily incidence rate of new cases and mortality up to 60 days later. Timelli and Girardi (16), within the context of a retrospective study, also noted that the timing of lockdown measures is vital for the control of the spread, as regions with delayed implementation of lockdown measures had a higher peak of cases (382– 921cases/100,000 vs, 265cases/100,000 cases). Similarly, Lilleri et al. (17) pointed out that earlier implementation of lockdown measures, when the number of cases is still low, might contribute to the flattening of the pandemic curve.

**Table 1.** Study characteristics and results for modelling and non-modelling studies with data from Italy (n=8)

| First Author, Year | Type of Study | Time horizon | Intervention(s) | Comparator | Results |  |  |
| --- | --- | --- | --- | --- | --- | --- | --- |
|  |  |  |  |  | Cases | Deaths | Hospital/ICU admissions |
| Gatto et al. 2020 (11) | SEIR model | 21/2-25/3/2020 | 9/3/2020 lockdown in Lombardy and 15 other provinces which was extended to the whole of Italy on 11/3/2020 | Absence of SD measures | **Absence of SD measures**<br>$R_0 = 3.60$ (95% CI: 3.49 to 3.84)<br>**Lockdown as of March 25th, 2020**<br>↓ 45% (95% CI: 42 to 49%) in transmission<br>0.226 x 106 (median) averted cases (95% CI: 0.172 x 106 to 0.347 x 106) | | |
| Palladino et al. 2020 (12) | Modelling study (undefined) | 24/2-3/5/2020 | 7-day earlier lockdown (2/3/2020) | Lockdown as implemented in Italy (9/3/2020) | **Lockdown from 9/3/2020**<br>210,717 total cases<br>**Lockdown from 2/3/2020**<br>84,626 total cases (-60%: -55% to -64%) | **Lockdown from 9/3/2020**<br>28,884 total deaths<br>**Lockdown from 2/3/2020**<br>16,080 total deaths (-44%: -38% to -50%) | **Lockdown from 9/3/2020**<br>105,359 non-ICU admissions<br>32,497 ICU admissions<br>**Lockdown from 2/3/2020**<br>50,644 non-ICU admissions (-52%: -46% to -57%)<br>16,885 ICU admissions (-48%: -42% to -53%) |
| Marzianoa et al. 2021 (13) | SIR model | up to 30/9/2020 | 1) Lifting of lockdown on 4/5/2020<br>2) Lifting of lockdown on 27/4/2020<br>3) Lifting of lockdown on 20/4/2020 | Lifting of lockdown on 18/5/2020 |  |  | **18/5/2020**<br>95,843 (63,597-140,269) cumulative hospitalised cases<br>**lockdown of 4/5/2020**<br>**With schools closed** 110,764 (71,637-168,761)<br>**With schools open** 514,625 (216,832-1,049,453)<br>**lockdown of 27/4/2020**<br>**With schools closed** 133,288 (83,118-217,441)<br>**With schools open** 693,107 (320,178-1,326,405)<br>**20/4/2020**<br>**With schools closed** 175,199 (100,967-303,521)<br>**With schools open** 835,260 (417,971-1,541,439) |
| Sjodin et al. 2020 (14) | SEIR model | 14-days period | 1) Medium adherence to community quarantine<br>2) Complete adherence to community quarantine | Non-compliance to community quarantine | **No adherence**<br>110 secondary infections<br>**Medium adherence**<br>70 secondary infections<br>**Complete adherence**<br>30 secondary infections |  |  |
| Silverio et al., 2020 (15) | Observational study (cohort) | up to 3/5/2020 | Lockdown implemented in Italy from 9/3/2020 | N/A | Positive correlation between confirmed cases before lockdown and mortality up to 60 days ($p < 0.001$, $R^2 = 0.57$) | | |
| Timelli et al., 2021 (16) | Observational study (cohort) | 24/2-11/5/2020 | Lockdown | N/A | Regions with delayed implementation of lockdown measures had a higher peak of cases (382–921 cases/100,000 vs. 265 cases/100,000 cases) |  |  |
| Lilleri et al., 2020<br>(17) | Observational<br>study (cohort) | up to 10/5/2020<br>(day 81) | Lockdown | N/A | An earlier lockdown, when cases are<br>still low (<5 cases/100,000 persons),<br>may have contributed to the flattening<br>of the pandemic curve. |  |  |

### Spain

#### Modelling studies

The SD measures implemented in Spain at the early stages of the pandemic were evaluated in two modelling studies. Hyafil et al. (18) predicted that R_0_ dropped from 5.89 (5.46-7.09 95%CI) to 1.86 (1.10-2.63 95%CI) after the declaration of a “state-of-emergency”, while a further decrease to 0.48 (0.15-1.17 95%CI) was noted after the implementation of a complete lockdown. With a similar SIR model, Casares & Khan (19) showed that the timing of implementation plays a significant role in COVID-19 spread. Specifically, while the SD measures initiated on March 14th, 2020 (including mobility restrictions, school closure, socioeconomic activity suspensions, and home confinement) led to profound reductions of approximately 89-93% in the accumulated number of infections, as well as on the number of deaths and hospitalisations, the results would have been strengthened by a 4-day earlier enforcement.

#### Non-Modelling studies

The only non-modelling study identified for Spain was a time-series analysis performed by **Santamaria & Hortal (20)**, which indicated an early generalized decrease in the reproduction number after the nationwide lockdown. In contrast, the strengthening of the lockdown only had a low further impact, and an increase in Rt was related to the loosening of the lockdown measures. These results indicated the significance of a more generalized lockdown for COVID-19 spread to be contained, as well as the limited impact of a stricter lockdown.

**Table 2.**
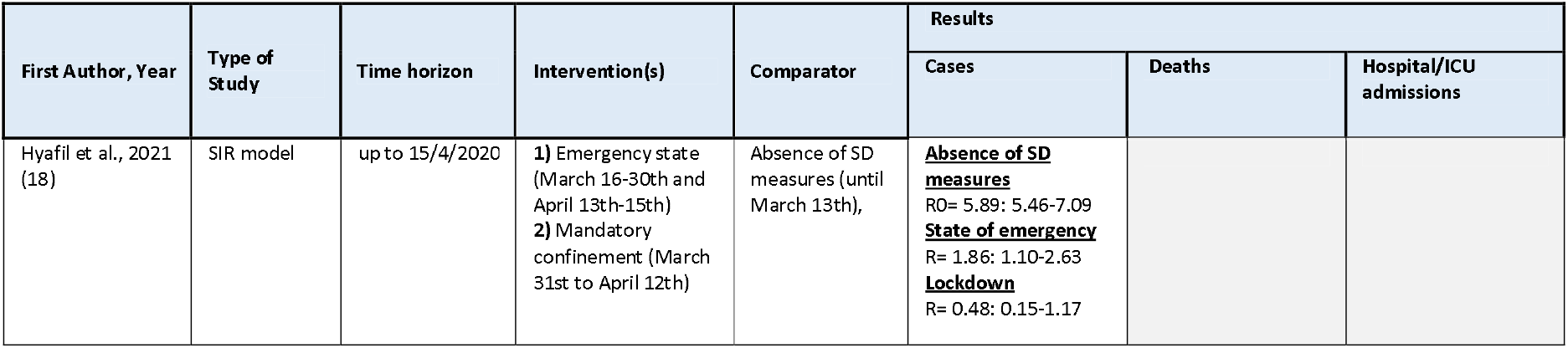
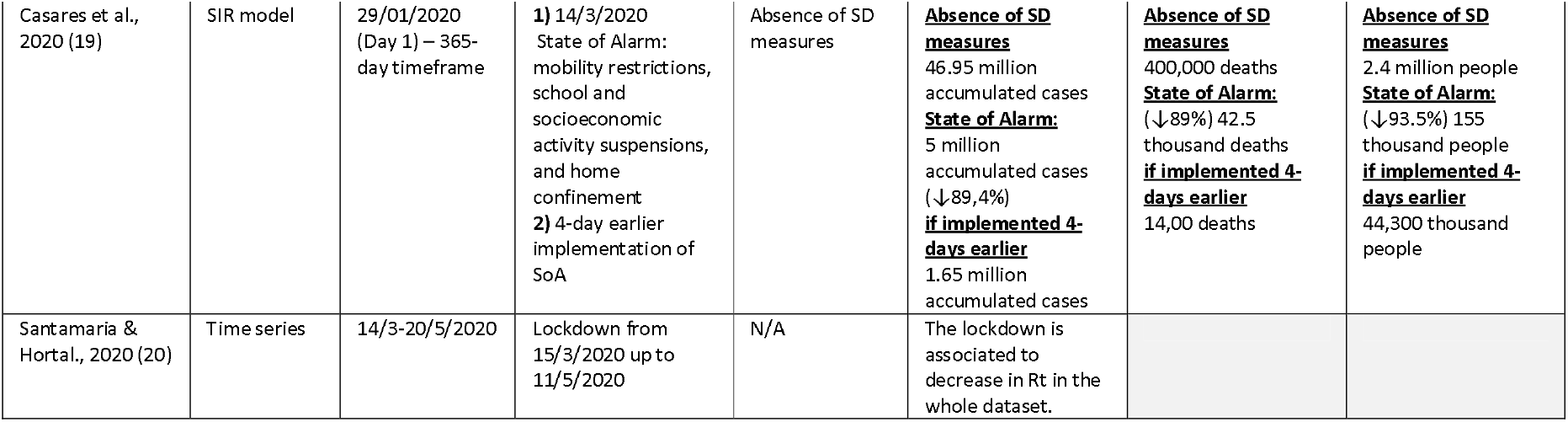
Study characteristics and results for modelling and non-modelling studies with data from Spain (n=3)

| First Author, Year | Type of Study | Time horizon | Intervention(s) | Comparator | Results |  |  |
| --- | --- | --- | --- | --- | --- | --- | --- |
|  |  |  |  |  | Cases | Deaths | Hospital/ICU admissions |
| Hyafil et al., 2021 (18) | SIR model | up to 15/4/2020 | 1) Emergency state (March 16-30th and April 13th-15th)<br>2) Mandatory confinement (March 31st to April 12th) | Absence of SD measures (until March 13th), | <u>Absence of SD measures</u><br>$R_0 = 5.89$: 5.46-7.09<br><u>State of emergency</u><br>$R = 1.86$: 1.10-2.63<br><u>Lockdown</u><br>$R = 0.48$: 0.15-1.17 | | |
| Casares et al., 2020 (19) | SIR model | 29/01/2020 (Day 1) – 365-day timeframe | 1) 14/3/2020 State of Alarm: mobility restrictions, school and socioeconomic activity suspensions, and home confinement<br>2) 4-day earlier implementation of SoA | Absence of SD measures | <u>Absence of SD measures</u><br>46.95 million accumulated cases<br><u>State of Alarm:</u><br>5 million accumulated cases (↓89,4%)<br><u>if implemented 4-days earlier</u><br>1.65 million accumulated cases | <u>Absence of SD measures</u><br>400,000 deaths<br><u>State of Alarm:</u><br>(↓89%) 42.5 thousand deaths<br><u>if implemented 4-days earlier</u><br>14,00 deaths | <u>Absence of SD measures</u><br>2.4 million people<br><u>State of Alarm:</u><br>(↓93.5%) 155 thousand people<br><u>if implemented 4-days earlier</u><br>44,300 thousand people |
| Santamaria & Hortal., 2020 (20) | Timeseries | 14/3-20/5/2020 | Lockdown from 15/3/2020 up to 11/5/2020 | N/A | The lockdown is associated to decrease in Rt in the whole dataset. |  |  |

### United Kingdom

#### Modelling studies

For the UK, five modelling studies were included in this review. Liu et al. (21) modelled the impact of the first 7-week national lockdown by using a Bayesian SEIR model to rebuild transmission dynamics. Estimations of Rt suggested that the SD measures, and particularly the implemented lockdown, effectively reduced COVID-19 transmissibility and curbed the COVID-19 pandemic as the Rt was generally <1 during the period of lockdown. A more detailed approach concerning the effectiveness of specific measures was followed by Rice et al. (22), who indicated that the largest number of deaths could be prevented with the combination of case isolation, voluntary quarantine of contacts and physical distancing of those aged over 70. Notably, when school closure was also added in this set of measures, the number of deaths was projected to increase, which according to the authors, might be attributed to the failure of prioritizing the protection of the vulnerable population. An assessment of different scenarios which could follow the lifting of lockdown on May 8th, 2020, was reported by Goscé et al. (23), who concluded that although an extended lockdown would be highly effective, other measures, including shielding of older people, mass testing and facemask use could all synergistically lead to a reduction of cases and deaths. The authors suggested, the most effective strategy seemed to be a combination of weekly mass testing, contact tracing and facemask use, in parallel with lockdown, which was projected to reduce deaths by 48% compared with lockdown alone. The lockdown measures implemented in the second COVID-19 wave were studied by Davies et al. (24), who adjusted an age-structured mathematical model to estimate the effect of different lockdown types implemented in Northern Ireland and Wales in October 2020, as well as to make projections for various epidemiological scenarios up to March 31st, 2021. The findings showed a reduction of 35% (30–41) in Rt, attributed to the Northern Ireland lockdown policy, and a 44% (37–49) decrease because of the lockdown in Wales, both with schools closed. Also, from October 1st, 2020, to March 31st, 2021, a projected COVID-19 epidemic would result in 280,000 hospital admissions and 58,500 deaths without SD restrictions, but these numbers could be reduced to 186,000 and 36,800, respectively, with a 4-week lockdown with schools remaining open. Closing schools was predicted to cause a further reduction in hospital admissions to 157,000 and deaths by 30,300. The authors concluded that a longer lockdown could reduce the number of deaths but would fail to reduce peak pressure on hospital services. Supportive findings were published by Yang et al. (25), who found that rolling interventions based on regional epidemiological data and with varying durations and intensities should be an effective strategy to control COVID⍰19 outbreaks in the UK. The authors suggested an intervention including suppression SD measures in London for 100 days and rolling interventions for three weeks in other regions to reduce the overall number of infections and deaths.

#### Non-Modelling studies

In England, the second wave was managed with progressive SD measures, which led to a national lockdown from November 5th to December 2nd, 2020, as presented in the study of Mensah et al. (2021) (26). According to the timelines, infections rates were maintained low in the early summer period, while an increase started from mid-August. The November lockdown contributed to the decrease in adult infection rates, followed by declines in student cases with one week lag. From November 23rd 2020, cases in both children and adults increased rapidly following the emergence of a more transmissible novel variant of concern. The beneficial effect of SD measures in England was also confirmed through the time-series data of Bernal et al. (27), who described the impact of physical distancing measures at week 13 of the COVID-19 pandemic. Timeline observations revealed a reduction in the number of outbreaks approximately three weeks after the implementation of NPIs. However, the number of outbreaks remained high through week 18, implying a possibly limited or delayed impact of NPIs in residential areas. A decline in hospital/ICU admissions and deaths started showing from weeks 14 and 15, respectively.

**Table 3.** Study characteristics and results for modelling and non-modelling studies with data from the UK (n=7)

| First Author, Year | Country, Area/ Population | Study type | Time horizon | Intervention(s) | Comparator | Results |  |  |
| --- | --- | --- | --- | --- | --- | --- | --- | --- |
|  |  |  |  |  |  | Cases | Deaths | Hospital/ICU admissions |
| Davies et al., 2021 (24) | England (Northern Ireland and Wales) | Age-structured mathematical model | 01/03-13/10/2020 (projections up to 31/3/2021) | **1) Northern Ireland's lockdown:** non-essential retail remained open, household bubbles of up to 10 people from 2 households<br>**2) Wales' lockdown:** non-essential retail was closed, stay at home, mixing with individuals from outside their households prohibited.<br>**3) School closure** | Absence of SD measures | **<u>Northern Ireland-type lockdown vs Absence of SD measures</u>**<br>**With schools closed:**<br>↓ 35% (30–41) in Rt<br>**With schools open:**<br>↓ 22% (15–27) in Rt<br>**<u>Wales-type lockdown vs Absence of SD measures</u>**<br>**With schools closed:**<br>↓ 44% (37–49) in Rt<br>**With schools open:**<br>↓ 32% (25–39) in Rt | **<u>Absence of SD measures</u>**<br>58,500 (55,800–61,100)<br>**<u>Northern Ireland</u>**<br>**With schools closed:**<br>34,900 (33,500–36,700)<br>**With schools open:**<br>41,500 (39,600–43,400)<br>**<u>Wales</u>**<br>**With schools closed:**<br>30,300 (29,000–31,900)<br>**With schools open:**<br>36,800 (34,900–38,800) | **<u>Absence of SD measures</u>**<br>280,000 (274,000–287,000)<br>**<u>Northern Ireland</u>**<br>**With schools closed:**<br>177,000 (171,000–181,000)<br>**With schools open:**<br>206,000 (199,000–213,000)<br>**<u>Wales</u>**<br>**With schools closed:**<br>157,000 (152,000–163,000)<br>**With schools open:**<br>186,000 (179,000–193,000) |
| Liu et al., 2021 (21) | England (9 regions) | SEIR model | 27/2-31/5/2020 | National lockdown from 23/3/2020 (lockdown studied period: 26/3-10/5/2020) | Before lockdown (control period: 12–26/3/2020) | **<u>Before lockdown</u>**<br>R0 between 2.8 and 3.9<br>**<u>After lockdown</u>**<br>Rt remained slightly <1 during most of the lockdown period |  |  |
| Rice et al., 2020 (22) | The UK and Northern Ireland | IBMIC model (Imperial's model) | The simulations are for 800 days, with day one being 1/1/2020. The simulated intervention period is for 91 days. | **1) Home case isolation**<br>**2) Voluntary home quarantine**<br>**3) SD for >70 years old**<br>**4) SD for the entire population**<br>**5) School and uni closure** | Different combinations of interventions |  | **<u>5</u>**<br>494-496 total deaths<br>**<u>1</u>**<br>416 total deaths<br>**<u>1+2</u>**<br>355 total deaths<br>**<u>1+2+4</u>**<br>411-440 total deaths<br>**<u>1+4</u>**<br>347-402 total deaths<br>**<u>1+2+3</u>**<br>261-262 total deaths<br>**<u>1+2+3+5</u>**<br>342-357 total deaths |  |
| Goscé et al., 2020 (23) | England, London | Modelling study | 30-day timeframe from 9/3/2020 | Post lockdown lifting scenarios (lockdown period: 23/3 - 8/5/2020):<br>1) Universal testing + less stringent SD measures<br>2) Shielding those > 60 years<br>3) Universal testing + face coverings use without a lockdown.<br>4) Universal testing, isolation of infectious cases and their contacts and use of face coverings during lockdown | Lift on 8/5 | **Lift on 8/5**<br>R0= 2.56<br>**<u>Universal testing (weekly-3 times/week) + less stringent SD measures</u>**<br>R= 1.87 – 2.07<br>**<u>Shielding those > 60 years</u>**<br>R= 3.07<br>**<u>Universal testing + face coverings use without a lockdown</u>**<br>R= 1.92<br>**<u>Universal testing, isolation of infectious cases and their contacts and use of face coverings during lockdown</u>**<br>R= 0.27 |  |  |
| Yang et al., 2021 (25) | UK |  | February-April 2020 | **1)** Suppression SD measures (lockdown from 23/3/2020)<br>**2)** Mitigation SD measures (delay phase from 12-23/3)<br>**3)** Rolling interventions | Before SD measures | **<u>Suppression SD measures (lockdown from 23/3/2020)</u>**<br>**Before 23/3:** R0= 2.73 [0.97–5.40]<br>**28/3:** Rt= 0.69 [0.59–0.79]<br>**<u>Mitigation SD measures (12-23/3)</u>**<br>**Before 23/3:** R0= 2.73 [0.97–5.40]<br>**27/5:** Rt= 0.98 [95% CI 0.88–1.09]<br>**<u>Rolling interventions</u>**<br>**Before 23/3:** R0= 2.73 [0.97–5.40]<br>**28/3:** Rt= 0.69 [0.59–0.79] |  |  |
| Mensah et al., 2021 (26) | England | Observational (time trend analysis) | July-December 2020 | Lockdown on 5/11/2020 (schools remained open) | N/A | The re-opening of schools after the half-term break (26/10/2020) was associated with a continuing increase in infection rates across all educational settings. In adults, trends in SARS-CoV-2 infection rates remained unchanged. Following national lockdown in November 05th 2020, cases in adults plateaued and then fell rapidly. Similar trends were observed in secondary and primary educational settings but with a one-week lag. |  |  |
| Bernal et al., 2021 (27) | England | Observational (time trend analysis) | From the start of the epidemic to week 18, 2020 | Mandatory social & physical distancing measures implemented from week 13 | N/A | Decline from week 16 | Decline from week 15 | Decline from end of week 14 both hospital and ICU admission rates |

### France

#### Modelling studies

For France, three modelling studies were included in this current review. The effect of lockdown as implemented in France (Île-de-France) was investigated by **Di Domenico et al. (28)** through a stochastic age-structured transmission model, concluding to an 81% reduction of the average number of contacts, which was projected to decrease R from 3.18 (3.09, 3.24) to 0.68 (0.66, 0.69). Further projections indicated that lifting lockdown without an exit strategy plan would unavoidably lead to a re-emergent situation. Moreover, the authors noted that the implementation of additional prolonged measures could delay the pandemic peak for at least two months and reduce the peak incidence by more than 80%, but would not manage to relieve the healthcare system from the pressure of hospital and ICU admissions. An age-targeted analysis considering lockdown measures in France was performed by **Roche et al. (29)**, who examined the implementation of lockdown strategies in particular age groups (i.e., 0-30, 30-60, >60), suggesting that either a complete lockdown or a partial one targeting young (0-30) and middle-aged adults (30-60) would be sufficient to achieve pandemic suppression, while indirectly decreasing mortality rates in older adults. They also found that a complete lockdown might prevent exhaustion in the healthcare system.

#### Non-Modelling studies

The SD measures implemented for the second COVID-19 wave were assessed by **Spaccaferri et al. (30)** in 22 metropolitan areas of France within the context of an observational study. A significant decrease in the incidence of COVID-19 confirmed cases, as well as in the number of hospital admissions from 7 to 10 days after the lockdown was in place, suggesting that the measures possibly exerted a positive impact.

**Table 4.** Study characteristics and results for modelling and non-modelling studies with data from France (n=3)

| Lead author, Year | Study type | Time horizon | Intervention(s) | Comparator | Results |
| --- | --- | --- | --- | --- | --- |
|  |  |  |  |  | Cases |
| Di Domenico et al., 2020 (28) | Stochastic age-structured transmission model | 17/03-11/05/2020 | Lockdown implemented in France between 17/03-11/05/2020 | Absence of SD measures | **Absence of SD measures**<br>R0= 3.18 (3.09, 3.24)<br>**Lockdown**<br>RLD= 0.68 (0.66, 0.69) |
| Roche et al., 2020 (29) | SEIR model | Undefined | 1) Lockdown for all age groups<br>2) Lockdown for those aged up to 30<br>3) Lockdown for those aged 30-60<br>4) Lockdown for those aged >60 | Absence of SD measures | A complete lockdown and a partial strategy targeting young and middle age classes were the only interventions that could achieve suppression. Focusing only on elderlies did not dramatically change the epidemic duration. |
| Spaccaferri et al., 2020 (30) | Observational (cohort) | up to 17/12/2020 | Curfew from 17/10/2020, lockdown from 30/10/2020, curfew from 15/12/2020 | N/A | The decrease was significant from 7 to 10 days after lockdown implementation --> - 15.7 to - 20.6% decrease in the incidence rate |

### Germany

#### Modelling studies

Aravindakshan et al. (31) studied the effectiveness of SD measures in Germany both from the perspectives of their implementation and lifting. Through a modified SEIR model, they found that if no SD measures were enforced, the number of cases would have a 24.6-fold (IQR: 20–29) rise. Moreover, if all SD measures and restrictions, taken into force between late February and early March, ended on April 21^st^, 2020, the daily number of cases would increase by 150% (IQR: 144– 156%) and could be reduced to 108% (IQR: 103.7–112.5%) with a 1-week delay. The predictions also indicated that lifting non-essential services closures would cause the minimum increase in daily cases, followed by lifting initial business closures. Overall, it was shown that the maintenance of some NPIs in place for an additional week was associated with a decrease in COVID-19 cases by up to 20%. The beneficial effect of SD measures introduced in Germany during the first wave was also highlighted in the modelling study of **Schlosser et al. (32)**, who used mobility data to show that lockdown had a significant effect on COVID-19 transmission by flattening the pandemic curve and slowing down the spread to regions with a geographical distance.

#### Non-Modelling studies

**Wieland (33)** conducted an interrupted time series to investigate the effectiveness of SD measures, which started from March 8th, 2020, with the cancellation of mass gatherings, followed by school closures on March 16th and a national lockdown announced March 23^rd^. Results showed a significant decrease of the daily growth rate from 22.8% (CI:22.4, 23.2]) before any measure took place to 6.6% (CI 05.9, 7.3) on March 10^th^. The second breakpoint was detected on March 26^th^ with a further reduction in daily growth from 6.8% to 1.9% (CI 1.7,2.0), and the third was noted on April 13^th^ when the daily growth rate shifted from 1.9% to 0.4% (CI 0.3, 0.4). Additionally, the loosening of measures did not lead to a re-emergence of cases during the study period.

**Table 5.** Study characteristics and results for modelling and non-modelling studies with data from Germany (n=3)

| Lead author, Year | Study type | Time horizon | Intervention(s) | Comparator | Results |
| --- | --- | --- | --- | --- | --- |
|  |  |  |  |  | Cases |
| Aravindakshan et al., 2020 (31) | SEIR model | 18/02-07/05/2020 and 21/04-19/07/2020 | 1) SD measures: contact restrictions, initial business closures, retail outlet closures, stay at home orders, non-essential business closures, closure of educational institutes, and border closures<br>2) Lifting SD measures | 1) Absence of SD measures<br>2) Keeping SD measures in place | **1) Absence of SD measures vs SD measures**<br>↑24.6-fold (IQR: 20–29) in cases<br>**2) Lifting SD measures vs keeping SD measures**<br>**Lifting contact restrictions**<br>*From 21/4:* ↑ 150% (IQR: 144–156%) in daily case numbers<br>*1 week delay:* ↑ 108% (IQR: 103.7–112.5%) in daily case numbers<br>**Lifting educational facilities closure**<br>*From 21/4:* ↑ 46.1% (IQR: 44.0–48.1%) in daily case numbers<br>*1 week delay:* ↑ 34.4% (IQR: 32.7–36.2%) in daily case numbers<br>**Opening of retail outlets**<br>*From 21/4:* ↑ 33.9% (IQR: 33.0–34.8%) in daily case numbers<br>*1 week delay:* ↑ 24.5% (IQR: 23.4–25.6%) in daily case numbers<br>**Lifting initial business closures**<br>*From 21/4:* ↑ 18.6% (IQR: 17.8–19.5%) in daily case numbers<br>*1 week delay:* ↑ 14.4% (IQR: 13.7–15.0%) in daily case numbers<br>**Lifting non-essential service closures**<br>*From 21/4:* ↑ 3.6% (IQR: 3.1–4.1%) in daily case numbers<br>*1 week delay:* ↑ 2.6% (IQR: 2.2–3.0%) in daily case numbers |
| Schlosser et al., 2020 (32) | SIR model | February-June 2020 | Lockdown | Before lockdown | The lockdown measures "flatten the curve" of the epidemic |
| Wieland et al., 2020 (33) | Time series | 15/2-31/5/2020 | 8/3/2020: cancellation of mass events, between 16-18/3: closure of schools<br>23/3: lockdown<br>20/4: ease of SD measures | N/A | **1st breakpoint on 10/3 (CI [March 09th, March 11th])**<br>Daily growth rate reduced from 22.8% (0.228, CI [0.224, 0.232]) to 6.6% (0.066, CI [0.059, 0.073]).<br>**2nd breakpoint on 26/3 (CI [March 25th, March 27th])**<br>Decrease in daily growth from 6.8% to 1.9% (0.019, CI [0.017, 0.020]).<br>**3rd breakpoint on 13/4 (CI [April 12th, April 14th])**<br>Daily growth rate shifted from 1.9% to 0.4% (0.004, CI [0.003, 0.004]) |

### Greece

#### Modelling studies

In Greece, the SD measures implemented for the management of the first pandemic wave were evaluated through a SEIR model designed by Sypsa et al. (34). According to the results, the Rt was projected to decrease by 42.7% after the closure of schools, shops and entertainment venues and by 81.0%, reaching 0.46, after the implementation of a national lockdown. The authors also made an attempt to delineate each measure’s impact concluding to an estimated reduction of approximately 1.1–1.3 in Rt if each measure was applied individually, and highlighted the significance of combining SD measures to strengthen the overall effectiveness.

**Table 1.**
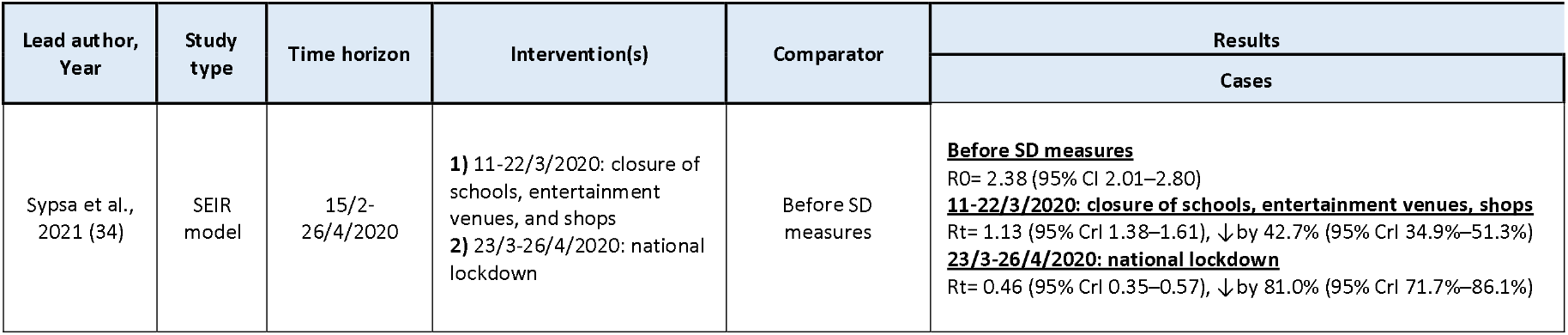
Study characteristics and results for modelling and non-modelling studies with data from Greece (n=1)

### Portugal

#### Modelling studies

In Portugal, a modelling study by **Ricoca Peixoto et al. (35)** forecasted 5568 averted cases, 146 averted deaths, and 519 averted ICU admissions between April 1^st^to 15^th^, 2020, after the lockdown implemented on March 16^th^, 2020. Among the contributing factors of the effectiveness of the SD measures, **Pais & Taveira (36)** focused on the level of population compliance and designed a simple SI model parameterized with data from the population of Portugal. Simulations showed that the benefit of pandemic mitigation increases with increasing population adherence to the control measures. However, as the authors stated, percentages of compliance over 75% might probably require a prolonged implementation period, which can result in economic/social burden, implying the multifactorial nature of SD measures.

**Table 7.** Study characteristics and results for modelling studies with data from Portugal (n=2)

| Lead Author, Year | Study type | Time horizon | Intervention(s) | Comparator | Results |  |  |
| --- | --- | --- | --- | --- | --- | --- | --- |
|  |  |  |  |  | Cases | Deaths | Hospital/ICU admissions |
| Ricoca Peixoto et al., 2020 (35) | ARIMA models (AutoRegressive Integrated Moving Average) | up to 15/4/2020 | Lockdown from 16/3/2020 | Absence of lockdown | **<u>Absence of lockdown</u>**<br>24,405 cases<br>**<u>After lockdown (1-15/4/2020)</u>**<br>5568 averted cases (-23%, n=18,837) | **<u>Absence of lockdown</u>**<br>588 deaths<br>**<u>After lockdown (1-15/4/2020)</u>**<br>146 averted deaths (-25%, n=442) | **<u>Absence of lockdown</u>**<br>748 ICU admissions<br>**<u>After lockdown (1-15/4/2020)</u>**<br>519 averted deaths (-69%, n=229) |
| Pais & Taveira et al., 2020 (36) | SI model | up to 19/8/2020 | SD measure compliance scenarios | Absence of measures | **<u>30-40% compliance</u>**<br>885,725–928,703 total cases<br>**<u>70-75% compliance</u>**<br>456,446–572,624 total cases<br>**<u>90% compliance</u>**<br>847 total cases | **<u>30-40% compliance</u>**<br>14,674–16,754 total deaths<br>**<u>70-75% compliance</u>**<br>5,288–9,056 total deaths<br>**<u>90% compliance</u>**<br>23 total deaths |  |

### Sweden

#### Modelling studies

Considering the COVID-19 situation in Sweden, **Sjodin et al. (37)** developed a SEIR model to examine different combinations of SD measures. The analysis revealed that the implementation of moderate SD measures in ages 0–59 years, alongside strong SD measures in ages over 60 years, and effective detection and isolation of infectious individuals would lead to the most significant reduction in infected cases and infection fatality rate. However, the authors concluded that although ICU demand could be maintained at non-overwhelming levels for the healthcare system with mitigation strategies, deaths are not able to be effectively prevented.

**Table 8.** Study characteristics and results for modelling studies with data from Sweden (n=1)

| Lead author, Year | Study type | Time horizon | Intervention(s) | Comparator | Results |  |
| --- | --- | --- | --- | --- | --- | --- |
|  |  |  |  |  | Cases | Deaths |
| Sjodin et al., 2020 (37) | SEIR model | 24/2-1/9/2020 | **1)** Modest SD in ages 0–59 years, moderate in ages >60 years<br>**2)** Modest SD in ages 0–59 years, moderately strong in ages >60 years<br>**3)** Moderate SD in ages 0–59 years, very strong in ages 60–79 years, strong in ages >80 years, and increased isolation of cases<br>**4)** Moderate SD in ages 0–59 years, strong in ages 60+ years, and further improved isolation of cases | Absence of SD measures | <u>**Absence of SD measures**</u><br>10,093,341 total cases<br>~~1~~ - 9,661,307 total cases<br>~~2~~ - 9,273,896 total cases<br>~~3~~ - 2,477,887 total cases<br>~~4~~ - 1,706,342 total cases | <u>**Absence of SD measures**</u><br>IFR= 0.46<br>~~1~~ IFR= 0.45<br>~~2~~ IFR= 0.42<br>~~3~~ IFR= 0.35<br>~~4~~ IFR= 0.30 |

### Denmark

#### Modelling studies

**Valentin et al. (38)** identified the basic reproduction number in Denmark before and after the lockdown enforcement with the design of a SEIR-type model, showing a significant reduction from 3.32 (95% PI: 3.31–3.33) to 0.92(95% PI: 0.92–0.93).

**Table 9.** Study characteristics and results for modelling studies with data from Denmark (n=1)

| Lead Author, Year | Country, Area/ Population | Study type | Time horizon | Intervention(s) | Comparator | Results |
| --- | --- | --- | --- | --- | --- | --- |
|  |  |  |  |  |  | Cases |
| Valentin et al., 2021 (38) | Denmark | SEIR model | 16/3-13/4/2020 | Lockdown from 18/3/2020 | Before lockdown | <u>**Before lockdown**</u><br>Ro= 3.32(95% PI: 3.31–3.33)<br><u>**28 days after lockdown**</u><br>Rt = 0.92(95% PI: 0.92–0.93) |

### Belgium

#### Non-Modelling studies

**Ingelbeen et al. (39)** assessed the influence of progressive lifting and re-enforcement of SD measures in Brussels during the second wave of the pandemic between August and November 2020. During the summer of 2020, an outburst in the number of cases in the second largest city forced strict physical distancing measures, which, however, were loosened after holidays despite the rising number of cases. From October 6th, 2020, SD measures started progressively to be re-introduced, and Rt, which peaked on September 17th, 2020, at 1.48 (95% CrI:1.35–1.63), decreased by 44.6% to 0.82 (95% CrI: 0.79–0.85) three weeks post to the closure of restaurants, bars, and sports facilities.

**Table 10.** Study characteristics and results for non-modelling studies with data from Belgium (n=1)

| Lead Author, Year | Country, Area/ Population | Study type | Time horizon | Intervention(s) | Comparator | Results |
| --- | --- | --- | --- | --- | --- | --- |
|  |  |  |  |  |  | Cases |
| Ingelbeen et al., 2021 (39) | Brussels | Observational study (time trend analysis) | August–November 2020 | From 6/8/2020, SD measures went into force progressively, including the closure of bars, cafes, restaurants and sports facilities, teleworking, restrictions in private gatherings, curfew | N/A | The $R_t$ =1.48 (95% CrI:1.35–1.63) on 17/9/2020. Three weeks after the closure of bars, restaurants and sports facilities, $R_t$ had decreased by 44.6% to 0.82 (95% CrI: 0.79–0.85) |

### Multiple Countries

#### Modelling studies

Consistently with the studies performed in each country, the reduction in COVID-19 transmission was also confirmed by **Gul et al. (40)** for Germany and Italy, who studied both countries up to April 16^th^, 2020. It was observed that with the adoption of strict interventions, there would be a reduction of R_0_ from 2.8-3.0 to 1.3-1.5. The beneficial effect of SD was also profound in the studies of **Bryant et al. (41)** and **Belloir et al. (42)**, who predicted reductions in Rt in all studied countries, except for Sweden and Denmark. Concerning the timing of implementation, **Palladino et al. (43)** indicated that the daily number of cases would have been reduced by 92%, 81%, 78% and 90% in France, Italy, Spain and the UK, respectively, if in each country the lockdown had been implemented earlier, three days after the first-50 cases, and not at the time of enactment.

#### Non-Modelling studies

A more generic analysis was performed by **Khataee et al. (44)**, who used data from nine European countries, including Italy, Spain, France, the UK, Germany, Switzerland, Netherlands, Belgium and Sweden, to quantitatively determine the impact of SD measures as implemented in each one of these countries through mobility data. The results indicated a drop in R_0_ in all studied countries, while a strong positive correlation between the decrease in the R and the mobility restrictions was also found. Moreover, the time between the peak of cases and the SD measures initiation was irreversibly associated with the stringency of measures. In more specific, the time from the national lockdown to the peak varied from 10 days in Italy to more than three weeks in Switzerland, whereas the time from mobility change to the peak ranged from 19 days in Italy and Spain up to 34 days in Sweden. Similar conclusions arise from the interrupted time series analysis of **Voko & Pitter (45)** in 28 European countries showing that the incidence of new COVID-19 cases, which grew by 24% on average daily before the change point, reduced in a range from 0.9% to 1.7% by increasing SD measures.

The impact of different lockdown durations was studied by **Coccia (46)** with a case series of six European countries, including Austria, France, Italy, Portugal, Spain, and Sweden, which were grouped under the categories of those with a 15-day duration of lockdown and those with a 61-day duration. Results suggested that countries with longer national lockdown duration had a higher fatality rate, while differences in COVID-19 cases were not significant. The authors attributed the increased fatality rate possibly to lower healthcare capacities and older populations forcing countries to maintain lockdown measures for more extended periods. Finally, **Martinez-Valero et al. (47)** indicated a significant direct correlation between the number of deaths and the time elapsed from the declaration of the initial case to the introduction of lockdown decision during the COVID-19 pandemic in 16 European countries.

**Table 12.**
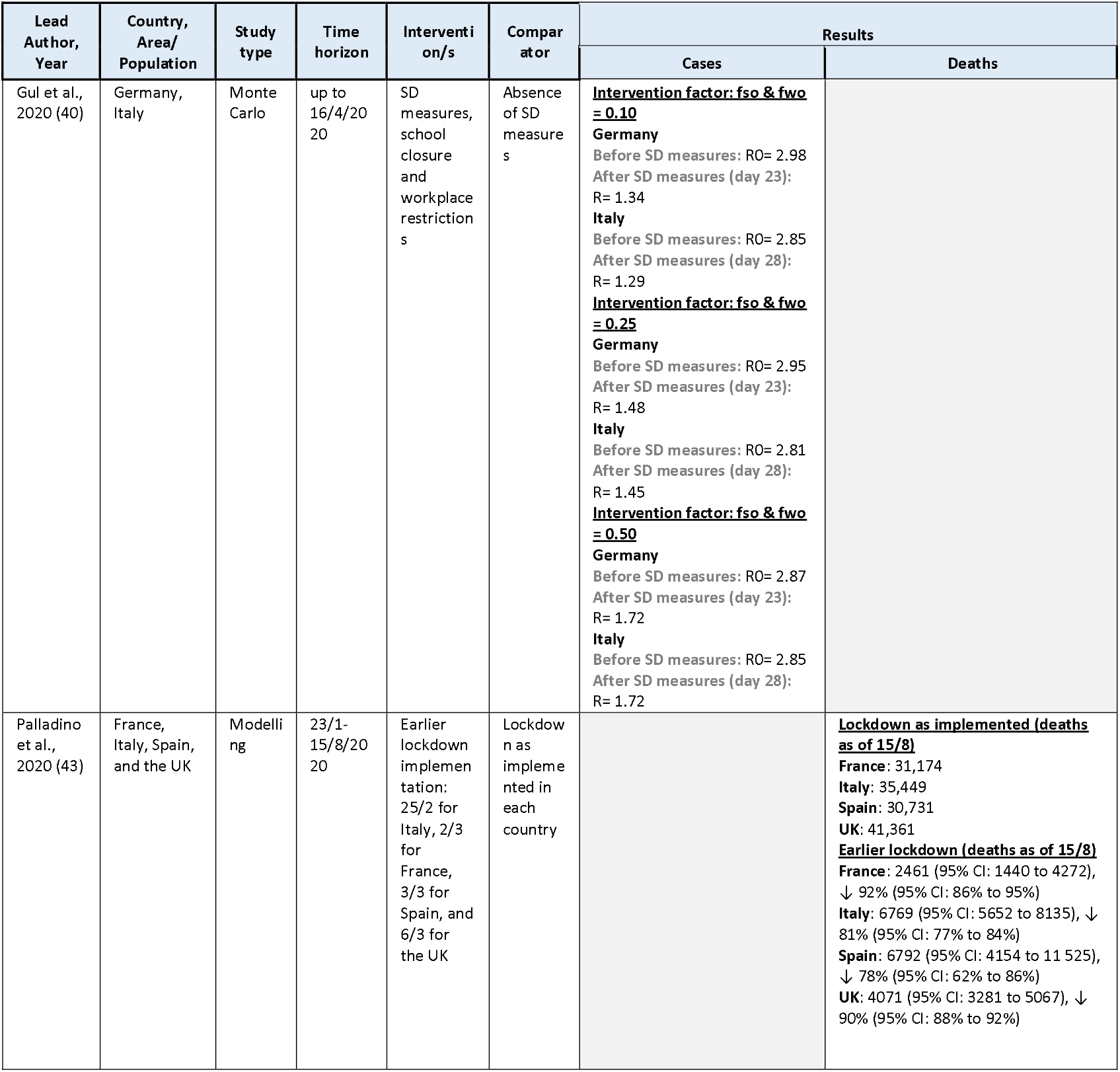
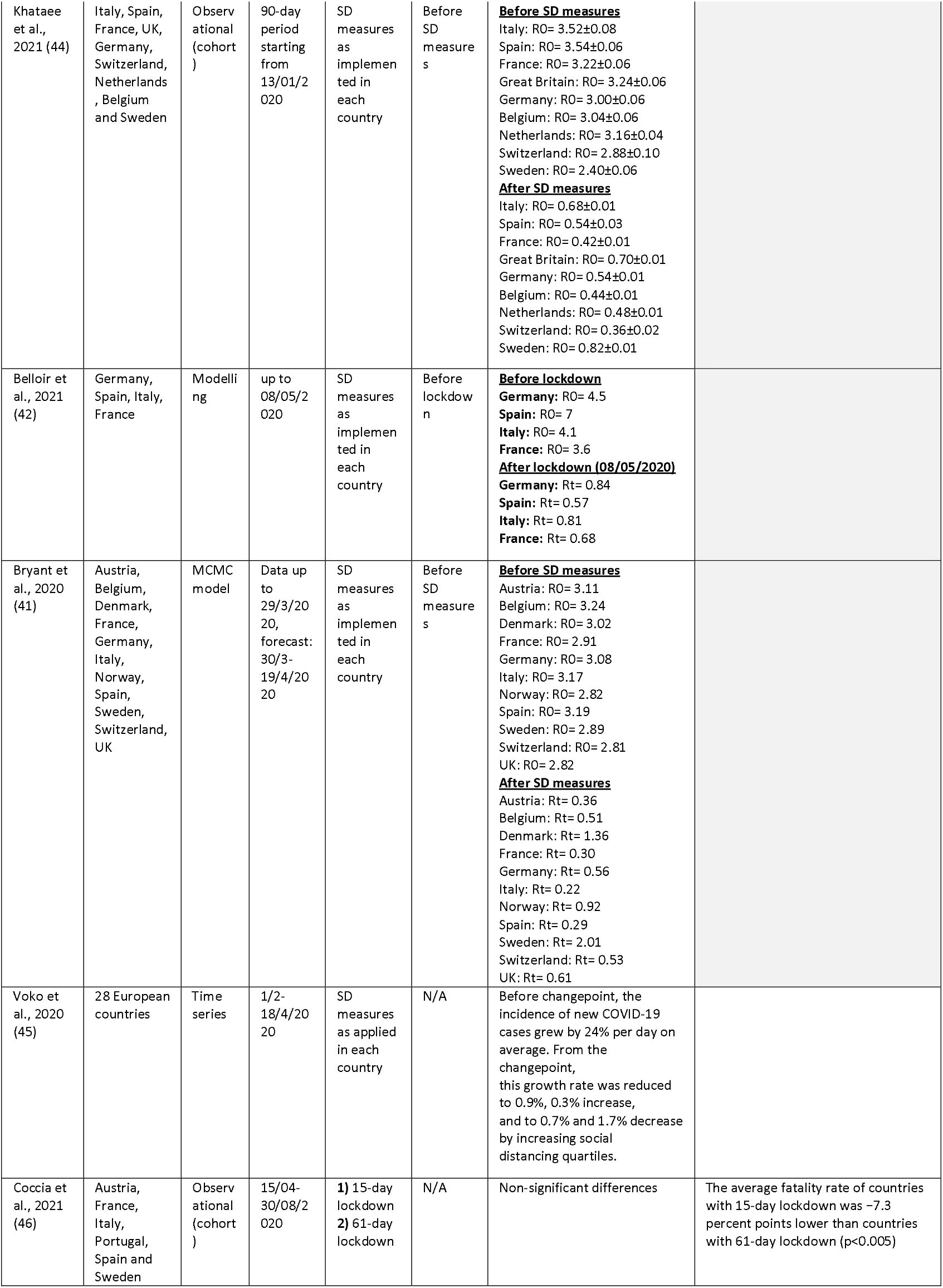
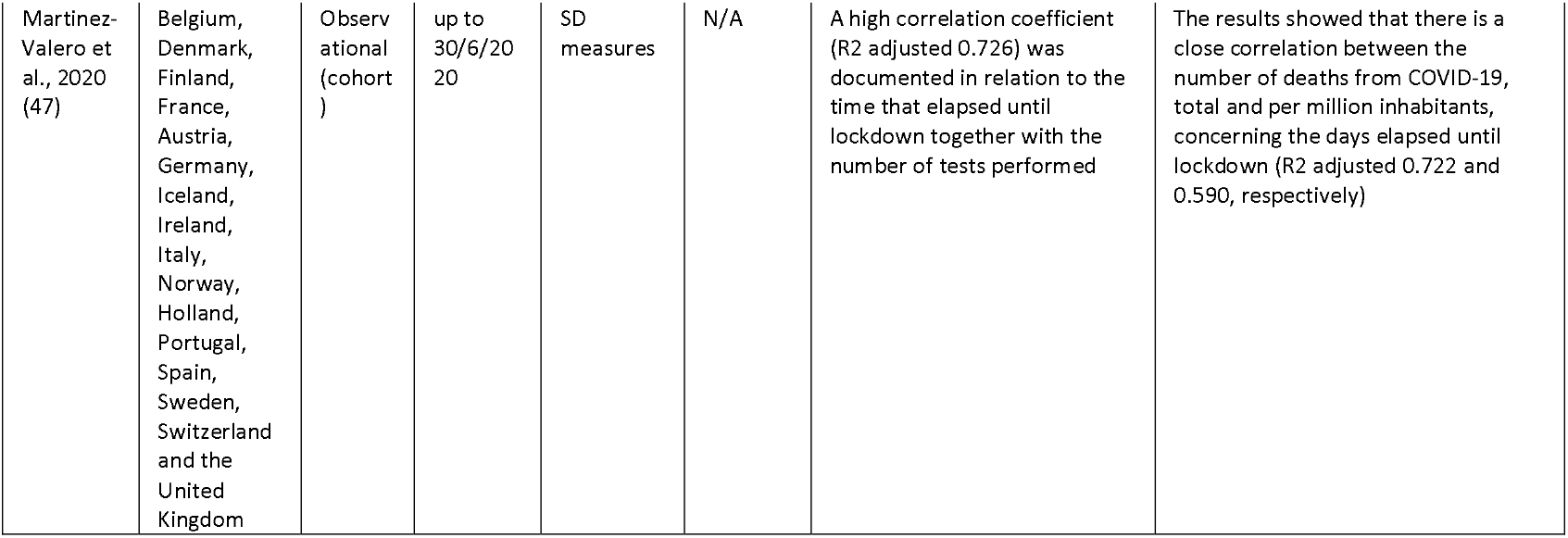
Study characteristics and results for modelling and non-modelling studies, which included data from multiple countries (n=6)

### School closure

#### Modelling studies

Among the 45 studies included in this current review, two examined in more depth the effectiveness of school closure as a separate measure. **Rozhnova et al. (48)** attempted to predict the impact of reducing school contacts in pandemic progression through an agent-structured transmission model with data from the Netherlands. The analysis showed that if complete school closure were implemented after the summer holidays in August 2020, R would be reduced by 10%, from 1.31 (95% 1.15–2.07) to 1.18 (95% 1.04–1.83). However, if school closure were enacted in November 2020, after implementing a partial lockdown since August, that would lead to a value of R of 1.00 (95% 0.94–1.33), it could further decrease R by 16%. Contact restrictions within the age group of 10-20 years old caused a slightly more significant reduction in Re compared to 5-10 years old. Notably, in the second wave of the pandemic, when primary and secondary schools remained open, their transmission role was limited. School closure was also assessed by **Rypdal et al. (49)** based on the population of two large cities of Norway, Oslo and Tromso, indicating that a controlled and gradual school re-opening would only have a slight increase in the reproduction number of less than 0.25, and probably in the range between 0.10 and 0.14, which would not substantially affect the infection rates.

**Table 13.** Study characteristics and results for modelling studies on the effectiveness of school closure (n=2)

| Authors | Country, Area/<br>Population | Study<br>type | Time horizon | Intervention(s) | Comparator | Results |
| --- | --- | --- | --- | --- | --- | --- |
|  |  |  |  |  |  | Cases |
| Rozhnova et al., 2021 (48) | Netherlands | Bayesian model | 27/02-30/04/2020 (hospital admission data)<br>April/May 2020 (seroprevalence data) | School closure in August and in November | Right before school closure | **August 2020**<br>Re = 1.31 (95% CrI 1.15–2.07)<br>**School closure in August**<br>Re (↓ 10%) = 1.18 (95% CrI 1.04–1.83)<br>**November 2020 (less stringent measures)**<br>Re = 1.00 (95% CrI 0.94–1.33)<br>**School closure in November**<br>Re (↓ 16%) = 0.84 (95% CrI 0.81–0.90)<br>**Reducing non-school-related contacts**<br>Re = 0.83 (95% CrI 0.75–1.10)<br>**School closure for 10–20-year-old children**<br>Re: ↓ 8%<br>**School closure for 5-10-year-old children**<br>Re: ↓ 5% |
| Rypdal et al. 2021 (49) | Norway (Oslo and Tromsø) | IBM model | up to 22/5/2020 | Kindergarten re-opening on 20/4/2020 and school re-opening on 27/4/2020 since the closure from 13/3/2020 | Before school re-opening | **After school re-opening**<br>Oslo: $\Delta R = 0.10$ (SE 0.004)<br>Tromsø: $\Delta R = 0.14$ (SE 0.005) |

### Case detection and management

Four modelling studies assessed the effectiveness of case detection and management strategies in this systematic review. **Willem et al. (50)** used the Simulate Transmission of Infectious Diseases (STRIDE) model with data from Belgium in order to contribute to the evidence on the effectiveness of contact tracing strategies. They found that the crucial timeframe for the completion of contact tracing during this period of the epidemic was four days after symptoms’ onset, while with a contact tracing strategy in place, an average decrease in hospital admissions of 22% in June and 57% in August could occur, assuming that at least 70% of the symptomatic cases are subjected to contact tracing and adhere to home isolation. **Almagor et al. (51)** developed an agent-based model based on the population of Glasgow, aiming at evaluating the effectiveness of a Contact-Tracing App (CTA). According to the results, the greatest reduction in overall and peak infections was noted when CTA was combined with a testing policy prioritising symptomatic cases. When CTA was adopted by 40-60% of the population and the testing capacity was between 1.5% and 3%, overall cases would decrease by 18-23% of the population and peak cases by 70-85%. In the case of 80% CTA adoption, the overall and peak cases would be reduced by an additional percentage of 7-12% and 4-19%, respectively. In the scenario where only COVID-19 tests were performed without any tracing, if 3% of the population was tested weekly, the overall number of infections could only decrease from 44 to 31% (by 13%), and the peak infections would reduce by 59%. A testing capacity of >3% did not lead to significant case reduction. The effectiveness of testing was also profound in the study of **Gul et al. (40)**, who modelled different testing ratios in Germany. In more specific, testing ratios of 1/1000, 5/1000, 1/100 and 5/100 of the population were found to cause a reduction in the mean number of deaths by 1%, 10%, 15%, 60%, respectively, compared to the no testing scenario. It could also be observed that random testing could be only considered effective if performed in over 5/1000 of the population. The authors noted that such testing numbers could be achieved regionally in smaller populations, but they may not be feasible for larger metropolitan cities. In the UK, **Panovska-Griffiths et al. (52)** also pointed out the significance of sufficient testing, tracing and isolating strategy to prevent a second COVID-19 wave during the autumn/winter of 2020. According to their model predictions, re-opening schools from September 1st, 2020, either full time or part-time with a rota system, along with the loosening of other SD measures, would induce a second pandemic wave in the absence of a scaled-up testing strategy. However, results showed that this might be prevented if a sufficient amount of symptomatic COVID-19 cases could be tested and an adequate number of their contacts could be traced. For example, in the case where 68% of contacts could be traced, approximately 65-75% of symptomatic cases would need to be tested and isolated, while if only 40% of contacts could be traced, the percentage of tested symptomatic cases would rise to 75-87%.

**Table 14.** Study characteristics and results for modelling studies on the effectiveness of case detection and management measures (n=4)

| Lead Author, Year | Country, Area/ Population | Study type | Time horizon | Intervention(s) | Comparator | Results |  |
| --- | --- | --- | --- | --- | --- | --- | --- |
|  |  |  |  |  |  | Cases | Deaths/Hospital |
| Almagor et al., 2020 (51) | Scotland, Glasgow/ Synthetic population of circa 103,000 agents from 2011 UK Census | An agent-based model | Undefined | Contact-tracing smartphone app (CTA) + COVID-19 detection tests | No CTA + testing | **<u>Testing without CTA vs. no CTA + testing</u>**<br>*3% testing capacity:* ↓13% overall infections, ↓59% peak infections<br>*>3% testing capacity:* no further decrease<br>**<u>CTA + testing prioritizing symptomatic cases vs. no CTA + testing</u>**<br>*40-60% CTA adoption + 1.5-3% testing capacity:* ↓18-23% overall infections, ↓70-85% peak infections<br>*80% CTA adoption + 1.5-3% testing capacity:* ↓30% overall infections, ↓89% peak infections<br>**<u>CTA with no priority testing of symptomatic cases vs. no CTA + testing</u>**<br>*40-80% CTA adoption + 3% testing capacity:* substantially more infections than the scenario with no CTA users |  |
| Gul et al., 2020 (40) | Germany | Monte Carlo model | up to 16/4/2020 | 1) Testing ratio of 1/1000 ppl<br>2) Testing ratio of 5/1000 ppl<br>3) Testing ratio of 1/100 ppl<br>4) Testing ratio of 5/100 ppl | No testing |  | **<u>1/1000 ppl vs no</u>**<br>↓ 1% of deaths<br>**<u>5/1000 ppl vs no</u>**<br>↓ 10% of deaths<br>**<u>1/100 ppl vs no</u>**<br>↓ 15% of deaths<br>**<u>5/100 ppl vs no</u>**<br>↓ 60% of deaths |
| Panovska-Griffiths et al., 2020 (52) | UK | Covasim model | 21/01-17/06/2020 | 1) 68% of contacts traced/no scale-up in testing<br>2) 68% of contacts are traced and testing is scaled up<br>3) 40% of contacts are traced and testing is scaled up | No testing | Across both scenarios of school re-opening the test-trace-isolate strategy would need to test a sufficiently large proportion of the symptomatic cases and trace their contacts with sufficiently large coverage, for R to diminish below 1 |  |
| Willem et al., 2021 (50) | Belgium | STRIDE model | Data up to 30/4/2020 and simulations up to 31/8/2020 | Contact tracing strategy | No contact-tracing |  | Average reduction in hospital admissions of 22% in June and 57% in August assuming that 70% of the symptomatic cases are subjected to contact tracing and comply with home isolation |

### Hygiene measures

Hygiene measures (i.e. facemasks) were only studied at the community level in this review, and hence one modelling study, one RCT and one quasi-experimental study were considered eligible.

#### Modelling studies

**Heald et al. (53)** developed a sequential assessment of the risk reduction provided by face coverings in public transport and retail shops in the UK using a step-by-step approach. They calculated the infection risk ratio and found a reduction of the risk score from 58.9 to 58.0 for a face covering efficacy of 20%, to 57.2 for a 40% efficacy, to 56.3 for an efficacy of 60%, and to 55.5 for an efficacy of 80%. A surgical mask, with an efficacy of over 90%, as used in hospitals, would reduce overall risk by 6.6% up to the maximum of 7.3%. The findings also showed that, with an R-value of 0.8 and face-covering of 40% effectiveness, average infections would be reduced by 844/week, hospital admissions by 8/week and deaths by 0.6/week. Nevertheless, If R was 1.0, the average community infections would remain at 29,400/week, and face coverings would reduce average weekly infections by 3,930, deaths by 2.9/week and hospital admissions by 36/week. Overall, the study showed that face-coverings, even with appropriate materials and maximum compliance on handling and wearing, produce limited benefits when used at low reinfection rates. Also, early implementation of such measured has a significant role in their effectiveness.

#### Non-Modelling studies

An RCT was performed by **Bundgaard et al. (54)** to evaluate the effectiveness of voluntary surgical face mask use in a sample of 4862 adults with no prior or current COVID-19 diagnosis or relevant symptoms who spent more than three hours out of home and have contact with other individuals. The results showed that 42 and 53 COVID-19 cases occurred in the intervention and control groups, respectively, without a statistically significant difference between them. As concluded, the recommendation of surgical mask use, supportively to other NPIs, did not decrease the infection rate by more than 50% among wearers in the context of a community with sufficient SD measures, modest infection rates, and uncommon general mask use.

The mandatory use of face masks was studied by **Mitze et al. (55)** in Germany, both in Jena and other regions. In Jena, the use of face masks in public transport and shops became mandatory before its compulsory implementation in all federal states, and within a 20-day period, it managed to reduce the number of newly registered cases by 75% relative compared to the control populations. Regarding the daily growth rate, it was reduced by approximately 70%, and however, when the analysis was performed in other regions, the effect was not as sizeable in the daily growth rate as it amounted to 14%, which rose, however, to 47% when only large cities were taken into consideration. The authors concluded that the timing of measures implementation may have played a crucial role in its effectiveness.

**Table 15.** Study characteristics and results for modelling and non-modelling studies on the effectiveness of case detection and management measures (n=3)

| Lead Author, Year | Country, Area/ Population | Study type | Time horizon | Intervention(s) | Comparator | Results |
| --- | --- | --- | --- | --- | --- | --- |
|  |  |  |  |  |  | Cases |
| Heald et al., 2021 (53) | UK | Modelling study (undefined model) | up to July 2020 | Mandatory mass use of face coverings on public transport from 15/6/2020, and in retail outlets from 24/7/2020 | No face mask | **Face mask 20%:** Infection Risk Score= 58 (↓ 1.5%)<br>**Face mask 40%:** Infection Risk Score= 57.2 (↓ 2.9%)<br>**Face mask 60%:** Infection Risk Score= 56.3 (↓ 4.4%)<br>**Face mask 80%:** Infection Risk Score= 55.5 (↓ 5.8%) |
| Bundgaard et al., 2020 (54) | Denmark/ 4,862 adults without COVID-19 who spend >3 hours per day outside home with exposure to other people | Experimental study (Two-arm, unblinded, randomised controlled trial) | April-May 2020 | SD measures + face mask (surgical) recommendation outside home | SD measures + no face mask (surgical) recommendation | **Intervention group**<br>42 participants<br>**Control group**<br>53 control participants<br>*Between-group difference* = - 0.3 percentage point (95% CI, - 1.2 to 0.4 percentage point; P= 0.38)<br>*OR* = 0.82 [CI, 0.54 to 1.23]; P= 0.33). |
| Mitze et al., 2020 (55) | Germany, Jena | Quasi-experimental study | April 2020 (20-days intervention period) | Mandatory mass use of face masks in public transport and shops from 6/4/2020 | Before mask use | **20-day mandatory mask use in public transport and shops vs. control**<br>**Jena**<br>↓ 23% in cumulative cases<br>↓ 75% in newly registered cases<br>↓ 70% in daily growth rate<br>**Other regions**<br>↓ 8.9% in cumulative cases<br>↓ 51.2% in newly registered cases<br>↓ 14% percentage points in daily growth rate |

## DISCUSSION

This systematic review aimed to update the evidence on the effectiveness of NPIs implemented at the meso and macro level to curb the COVID-19 pandemic in the European Region. Given the pandemic’s sudden escalation, the rapidly rising numbers of cases and deaths and the exhaustion of healthcare systems, policymakers had to make timely decisions on the type of strategies to be adopted with only limited evidence-informed guidance, especially in the early pre-vaccine stages of the pandemic which are primarily reflected within this review. The literature presented here predominantly reports on research undertaken during 2020, thus offering a comprehensive assessment of the effectiveness of NPIs in Europe during the first and second waves of the COVID-19 pandemic, in spring and autumn 2020, respectively.

### Physical distancing measures

Physical distancing measures, including the closure of educational institutions, entertainment venues, non-essential businesses, gyms and sports, workplace closure, bans on gatherings, cancellation of events, isolation of cases and quarantine of contacts, and stay-at-home recommendations/orders were a core component of the control strategies for the containment of COVID-19 pandemic in most of the studied countries. In this current systematic review, most studies attempted to investigate various combinations of physical distancing measures, with lockdown strategies gaining the most attention. Overall, both empirical data and modelling studies showed that physical distancing measures slowed COVID-19 spread and reduced hospital/ICU admissions and mortality in all EU/UK/EEA countries included in this review. The individual effect of individual NPIs is difficult to be estimated due to their parallel implementation and the multifactorial nature that affects their impact, both within and across countries.

A significant overarching factor that was noted to impact physical distancing measures’ effectiveness is the ***timing*** of the implementation, a factor noted by both modelling and observational studies. Palladino et al. (12) predicted that a 7-day earlier lockdown introduction would have averted a significant number of cases, deaths, hospital and ICU admissions in Italy. Moreover, it was modelled that the daily number of cases would have been reduced by 78-92% in France, Italy, Spain and the UK, respectively, if in each country the lockdown would have started three days after the first 50 cases, and not the actual days of enactment (43). Consistent were the results of Casares & Khan (19), who showed with a SIR model that the timing of implementation plays a significant role in COVID-19 spread. The positive impact of an earlier performance of lockdown measures was also profound in the non-modelling studies performed in Italy that indicated that regions with a delayed lockdown implementation had a peak with a higher number of cases than those with a shorter implementation delay. (16) (16)

In addition to timing, the ***duration*** of implementation is also a significant aspect that has to be taken into consideration in the decision-making process. In general, the results of the studies included in this systematic review used simulation models that differ in design, baseline parameters and assumptions; however, they indicated that an earlier lifting of SD measures would lead to an increase in cases in Germany (31) and higher hospitalisation rates in Italy (13). Goscé et al. (23) concluded that an extended lockdown for two-three more months would be the most effective, although other measures including shielding of older people, mass testing and facemask use could all synergistically lead to a reduction of cases and deaths. A recent systematic review, published in April 2020, noted that findings consistently indicate that quarantine measures are important in reducing incidence and mortality during the COVID-19 pandemic, although there is uncertainty over the magnitude of the effect (56). Although lockdowns have an impact on SARS-CoV-2 transmission, the duration itself depends on many factors, such as the COVID-19 spread at the time of control initiation, the population’s level of compliance, and the capacity of the healthcare system. At the same time, as they also have an impact on population wellbeing (57, 58) and the economy (46), a fine balance is needed between mitigating the pandemic and promoting economic and social wellbeing.

Another critical factor noted by multiple authors to highly affect the effectiveness of NPIs is the level of acceptance, perceived effectiveness, and compliance from the public, which, however, were not the focus of this specific review. Only one study within our review (36) focused on the level of population compliance with simulations showing that the benefit in pandemic mitigation increases with the increase of population adherence to the control measures. Data collected on March 2020, across the G7 countries showed that those who were concerned about the impact of COVID-19 on their own or their children’s education or on their personal income were more likely to practice personal protective measures and physical distancing (59).

As for school closure, the two included modelling studies indicated only a moderate effect on COVID-19 cases when implemented individually. At the same time, if appropriate measures were applied systematically in school premises, R would not be significantly affected. Similar conclusions were presented in the rapid systematic review of Viner et al., who underlined that the only study examining school closures independently found relatively marginal impact by preventing only 2-4% of deaths, compared to other physical distancing interventions (60). Considering transmission, in the review conducted by Suk et al. children were most frequently transmitting SARS-CoV-2 in household settings, while examples of children as index cases in school settings were rare (61).

### Case detection and management and hygiene measures

The evidence on the effectiveness of case detection and management strategies is limited due to the lack of empirical data within this review, as only four modelling studies on this topic were included. Overall, the significant contribution of the timely isolation of a high proportion of cases for an adequate duration in limiting the SARS-CoV-2 spread, combined with contact tracing and quarantine of contacts, was highlighted in all studies. However, given the modelling design, the actual effect of the case detection and management measures as implemented in each country has not been captured along with the contributing factors, among which the availability of tests has a crucial role.

With regards to hygiene measures, only three studies were included in this review evaluating the effectiveness of face masks with different methodological designs in Europe at the meso and macro level (53-55). In general, the results indicated that they have a supportive effect in mitigating the pandemic spread. However, these data reflect on the early period of the pandemic when people were not as familiar with personal protective measures as on the later stages.

Despite the lack of studies that have assessed the impact of facemasks at the community and population level, there is a plethora of evidence that indicate the role of facemasks to mitigate respiratory pandemics, both as a cost effective preventive measure (3) and as a mitigation measure for COVID-19 transmission (62, 63).

Given that the studies included in this current review are heterogeneous regarding models, setting, timeframe and NPI interventions assessed, a direct comparison of the same class of NPIs in different countries should be performed with extreme caution. Discrepancies were noted in different countries even in the implementation of high visibility measures, like the stay-at-home orders. Additionally, the same national measures were implemented in a very different way at different times within the same country, with stay-at-home orders in spring 2020 being much stricter than in the autumn of the same year. One study that attempted to rank the effectiveness of COVID-19 interventions worldwide noted that the performance of all NPIs depends highly on the geographical region and the socioeconomic context, while accountability to government and political stability were found to exert influence(64). A more recent review of only empirical studies worldwide conducted by Mendez-Brito et al. (65) indicated that school closure, followed by workplace and entertainment venue closure, as well as bans of public events were the most effective NPIs, concluding that an early response and a combination of specific physical distancing measures are of crucial importance for the reduction of COVID-19 cases and deaths.

This report is part of the ECDC work related to collecting and analysing information on NPIs implemented for the COVID-19 pandemic. Within this frame, information on NPIs implemented in the EU/EEA since 1^st^ January 2020 is gathered in the ECDC-JRC Response Measures Database, which is publicly available at https://covid-statistics.jrc.ec.europa.eu/RMeasures.

### Strengths and limitations

Our systematic review has several strengths, including the systematic approach during study identification, data extraction and quality appraisal that was applied. Some limitations should also be acknowledged. We assessed peer-reviewed and published evidence available until April 2021, much of which included the period before December 2020, and hence the information provided does not adequately cover NPIs in place during the circulation of all variants of concern. As per our inclusion criteria, only studies based in EU/EEA and UK were assessed, and accordingly, the results may not reflect the effectiveness of NPI implementation in other contexts globally – although other systematic reviews of a broader scope indicate similar findings (66, 67). Furthermore, a significant percentage of identified studies were based on mathematical modelling studies that require many assumptions related to the model design, which may lead to variability in model predictions and uncertainty for decision-makers. There was, furthermore, significant heterogeneity in study designs and studied interventions/combination of interventions, which only allowed for a narrative presentation of the results. Also, given that NPI effectiveness depends on the implementation, the personal protective measures that are probably implemented in parallel, demographic characteristics, and compliance levels, direct comparisons across countries and across NPIs were not possible.

## Conclusions

It will continue to be important to refine understandings of the effectiveness of NPIs for controlling COVID-19, and the evidence generated during the COVID-19 pandemic may also be applicable to future pandemics. The findings of this review provide a narrative synthesis of the literature available for the European Region and could also be the basis for the development of a future guiding framework, including best practices and approaches to policymakers primarily in the EU and EEA countries. In conclusion, NPIs, as assessed within the context of this systematic review at the macro and meso level, are effective in reducing transmission rates, hospitalisation rates and deaths in the European Region and may be applied as response strategies to reduce the burden of COVID-19 in forthcoming waves as well as in other respiratory viruses and future outbreaks. However, particular attention should be paid on the timing and duration of these measures in order maximum benefits to be achieved.

## Data Availability

No new data have been produced in the present work

## ACKNOWLEDGEMENTS

We would like to thank Chrysa Chatzopoulou and Katerina Papathanasaki for contributing to data archiving and management. We would also like to thank Rene Niehus for his helpful comments on an earlier version of this work.

This report was commissioned by the European Centre for Disease Prevention and Control (ECDC), to the PREP-EU Consortium, coordinated by the School of Medicine, University of Crete, under specific contract No.7 ECD. 23661 within Framework contract ECDC/2019/001 Lot 1B.

## Disclaimer

This report was produced under a service contract with the European Centre for Disease Prevention and Control (ECDC), acting under the mandate from the European Commission. The information and views set out in this piece of work are those of the authors and do not necessarily reflect the official opinion of the Commission/Agency. The Commission/Agency do not guarantee the accuracy of the data included in this analysis. Neither the Commission/Agency nor any person acting on the Commission’s/Agency’s behalf may be held responsible for the use which may be made of the information contained therein.

## DATA AVAILABILITY STATEMENT

Data sharing not applicable as no datasets generated and/or analysed for this study.

## ETHICS STATEMENT

For the purposes of this review publicly accessible documents were used as evidence, and, hence, no ethics approval was required.

