## Supplementary Material for "Effectiveness of non-pharmaceutical measures (NPIs) on COVID-19 in Europe: A systematic literature review"

### Supplement 1: Search strategies for identifying studies

Database: Ovid MEDLINE(R) ALL <1946 to April 14, 2021>

| **KEY TERMS** | **HITS** |
| --- | --- |
| 1 exp Coronavirus/ | 67239 |
| 2 exp Coronavirus Infections/ | 81178 |
| 3 (Coronavir* or nCov or covid or covid-19 or Middle East Respiratory Syndrome or MERS or Severe Acute Respiratory Syndrome or SARS).ti,ab,kf. | 142566 |
| 4 1 or 2 or 3 | 149765 |
| 5 child day care/ | 5709 |
| 6 Schools/ or Universities/ | 82781 |
| 7 Nurseries, Infant/ | 1090 |
| 8 (school$ or nurser$ or pre?school$ or pre school or kindergarten or day care or daycare or child or infant).ti,ab,kf. | 882615 |
| 9 (Closure or close or closing or close?down or lock?down or shut?down closedown or lockdown or shutdown).ti,ab,kf. | 467942 |
| 10 5 or 6 or 7 or 8 | 927070 |
| 11 9 and 10 | 15494 |
| 12 exp Personal Protective Equipment/ or masks/ or protective devices/ or personal protective equipment/ or respiratory protective devices/ or Eye Protective Devices/ | 38816 |
| 13 (Mask? or facemask? or face-mask? or ppe or ipc or N95 or ffp or ffp1 or ffp3 or ffp2 or (filter* adj face adj piece) or ((face or respiratory or eye) adj2 (shield or equipment? or protect* or cover*)) or ((airborne or air-borne or droplet*) adj precau*) or N99 or N97 or respirator? or goggle? or ((safety or protective) adj (supply or supplies or device* or equipment? or material* or measure* or gear?)) or (safely adj1 equipped) or ((head or face) adj cover?) or ((physical or person*) adj (intervention* or barrier? or protect*)) or (protective adj clothing?)).ti,ab,kf. | 554157 |
| 14 12 or 13 | 577399 |
| 15 Travel by Air/ | 440 |
| 16 (travel ban$ or travel restriction$ or public transport or train$ or bus or buses or harbour or harbor or border crossing$ or travel advice or travel guidance or border scanning or (clos$ adj1 borders)).ti,ab,kf. | 624038 |
| 17 15 or 16 | 624450 |
| 18 Social Distance/ | 0 |
| 19 (physical distanc$ or social distanc$ or close contact$ or ((patient? or person* or individual?) adj1 isolat*) or distanc* or space or spacing or separation or meter? or metre? or foot or feet or transmission*).ti,ab,kf. | 1364883 |
| 20 18 or 19 | 1364883 |
| 21 ((((non-pharm* adj intervention*) or community intervention or stay at home or business clos$ or clos$) adj2 business) or (public gather adj2 ban) or lock?down or work from home).ti,ab,kf. | 7037 |
| 22 (randomized controlled trial or controlled clinical trial or multicenter study or pragmatic clinical trial).pt. or (randomis* or randomiz* or randomly).ti,ab. or groups.ab. or (trial or multicenter or multi center or multicentre or multi centre).ti. or (intervention? or effect? or impact? or controlled or control group? or (before adj5 after) or (pre adj5 post) or ((pretest or pre test) and (posttest or post test)) or quasiexperiment* or quasi experiment* or pseudo experiment* or pseudoexperiment* or evaluat* or time series or time point? or repeated measur*).ti,ab. | 11349554 |
| 23 Non-Randomized Controlled Trials as Topic/ | 895 |
| 24 interrupted time series analysis/ | 1187 |
| 25 Controlled Before-After Studies/ | 604 |
| 26 22 or 23 or 24 or 25 | 11349676 |
| 27 11 or 14 or 17 or 20 or 21 | 2489383 |
| 28 4 and 27 | 57413 |
| 29 28 and 26 | 26925 |
| 30 humans.sh. | 19160315 |
| 31 29 and 30 | 15134 |
| 32 limit 31 to yr="2020-Current" | 12846 |
| 33 limit 32 to english language | 12470 |

Database: Embase <1974 to 2021 April 14>

| **KEY TERMS** | **HITS** |
| --- | --- |
| 1 exp coronavirus/ | 33880 |
| 2 exp coronavirus infections/ | 113500 |
| 3 (Coronavir* or nCov or covid or Middle East Respiratory Syndrome or MERS or Severe Acute Respiratory Syndrome or SARS).ti,ab,tw. | 140700 |
| 4 1 or 2 or 3 | 153220 |
| 5 Schools/ or Universities/ or Child Day Care Centers/ | 140292 |
| 6 (schools or nursery or nurseries or preschool* or pre school or kindergarten or day care or daycare).ti,ab,kw. | 168926 |
| 7 (closure or close or closing or close?down or lock?down or shut?down or lockdown or shutdown or closedown).ti,ab,kw. | 583100 |
| 8 (5 or 6) and 7 | 5310 |
| 9 'mask'/de or 'protective equipment'/de or 'respiratory protection'/de or 'eye mask'/de or goggles.ti,ab,kw. | 1784 |
| 10 (mask* or facemask* or 'face mask' or ppe).ti,ab,kw. | 113257 |
| 11 ((filter* adj face adj piece) or ((face or respiratory or eye) adj2 (shield or equipment* or protect* or cover*))).ti,ab,kw. | 6217 |
| 12 (((safety or protective) adj (supply or supplies or device* or equipment* or material* or measure* or gear*)) or (safely adj1 equipped)).ti,ab,kw | . 22855 |
| 13 9 or 10 or 11 or 12 | 138568 |
| 14 (travel ban* or travel restriction or public transport or train* or bus or buses or harbour or harbor or border crossing or travel advice or travel guidance or border scanning).ti,ab,kw. | 848224 |
| 15 (distanc* or space or spacing or separation or physical distanc* or social distanc* or close contact* or ((patient* or person* or individual*) adj 1isolat*)).ti,ab,kw. | 976726 |
| 16 (meter* or metre* or foot or feet or ('non pharm*' adj intervention*) or ((physical or person*) adj (intervention* or barrier* or protect*)) or transmission*).ti,ab,kw. | 669963 |
| 17 14 or 15 or 16 | 2397327 |
| 18 (travel ban* or travel restriction or public transport or train* or bus or buses or harbour or harbor or border crossing or travel advice or travel guidance or border scanning).ti,ab,kw. | 848224 |
| 19 (clos* adj border*).ti,ab,kw. | 53 |
| 20 18 or 19 | 848274 |
| 21 (closure or close or closing or close?down or lock?down or shut?down or lockdown or shutdown or closedown).ti,ab,kw. | 583100 |
| 22 ((Non-pharm* adj intervention) or community intervention or stay at home or business clos*).ti,ab,kw. | 4320 |
| 23 (clos* adj2 business).ti,ab,kw. | 83 |
| 24 (public gather* adj2 ban*).ti,ab,kw. | 9 |
| 25 randomized controlled trial/ or controlled clinical trial/ or quasi experimental study/ or pretest posttest control group design/ or time series analysis/ or experimental design/ or multicenter study/ or (randomis* or randomiz* or randomly).ti,ab. or groups.ab. or (trial or multicentre or multicenter or multi centre or multi center).ti. or (intervention? or effect? or impact? or controlled or control group? or (before adj5 after) or (pre adj5 post) or ((pretest or pre test) and (posttest or post test)) or quasiexperiment* or quasi experiment* or pseudo experiment* or pseudoexperiment* or evaluat* or time series or time point? or repeated measur*).ti,ab. | 14735048 |
| 26 8 or 13 or 17 or 20 or 21 or 22 or 23 or 24 | 3011384 |
| 27 26 and 4 and 25 | 16020 |
| 28 limit 27 to yr="2020-Current" | 14609 |
| 29 limit 28 to english language | 14306 |

Cochrane Database of Systematic Reviews (15 April 2021)

1 Covid
2 Travel or closure or personal protective or distanc*
3 1 and 2 (17 hits)

Evidence Aid via Campbell Database of Systematic Reviews (15 April 2021)-

Preventing infection and transmission

Community level interventions

(16 hits)

### Supplement 2: Quality Appraisal of the included studies

Supplementary Table 1. Results from assessing the quality of individual modelling studies (n=30)

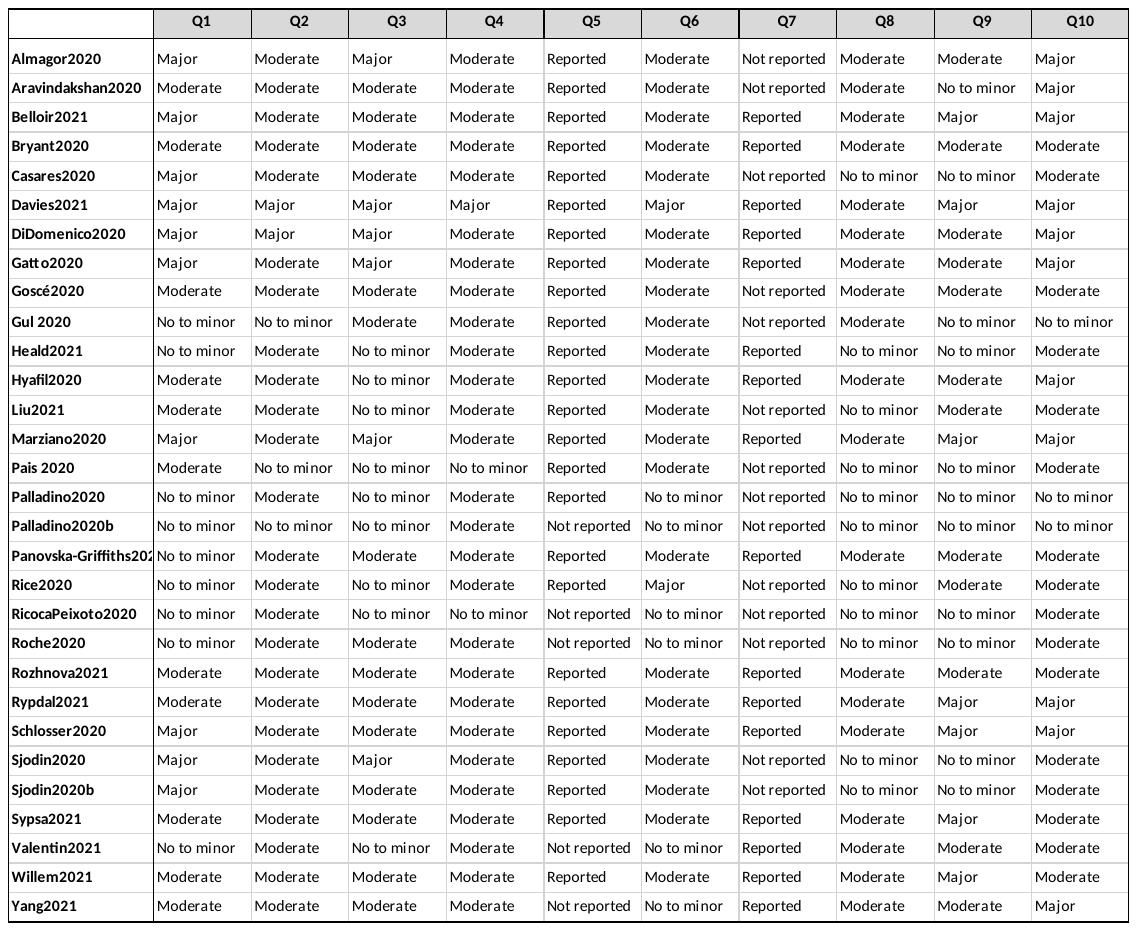

Supplementary Table 2. Results from the JBI critical appraisal tool for RCTs (n=1)

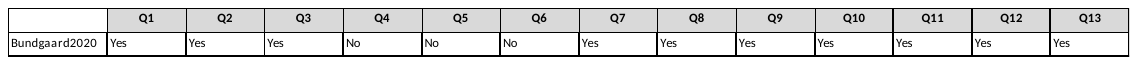

*Supplementary* *Table 3. Results from the JBI critical appraisal tool for quasi-experimental studies (n=1)*

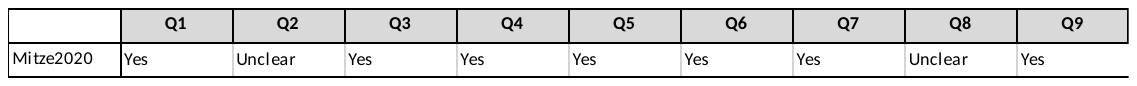

*Supplementary* *Table 4**. EPOC RoB for time-series studies (n=3)*

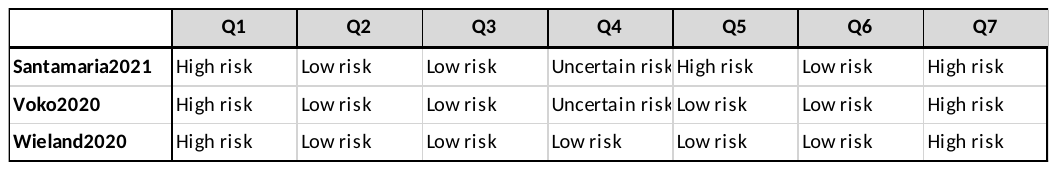

Supplementary Table 5. Results from the JBI critical appraisal tool for cohort studies (n=7)

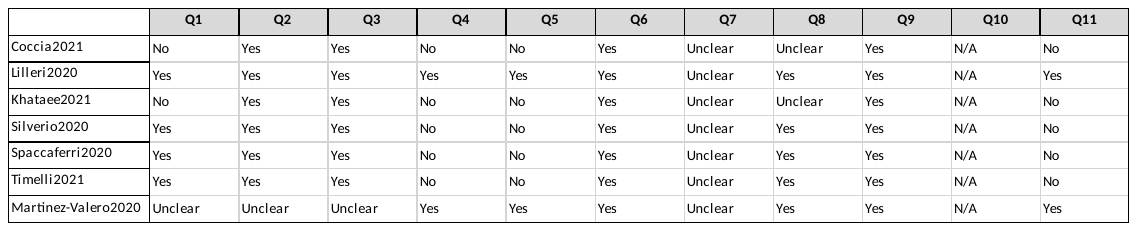
